# Short-Term Immune Response After Inactivated SARS-CoV-2 (CoronaVac®, Sinovac) And ChAdOx1 nCoV-19 (Vaxzevria®, Oxford-AstraZeneca) Vaccinations in Thai Health Care Workers

**DOI:** 10.1101/2021.08.27.21262721

**Authors:** Watsamon Jantarabenjakul, Napaporn Chantasrisawad, Thanyawee Puthanakit, Supaporn Wacharapluesadee, Nattiya Hirankarn, Vichaya Ruenjaiman, Leilani Paitoonpong, Gompol Suwanpimolkul, Pattama Torvorapanit, Rakchanok Pradit, Jiratchaya Sophonphan, Opass Putcharoen

## Abstract

**Background:** Inactivated SARS-CoV-2 (CoronaVac®,Sinovac, or SV) and ChAdOx1 nCoV-19 (Vaxzevria®,Oxford-Astra Zeneca, or AZ) vaccines have been administered to the health care workers (HCWs) in Thailand.

**Objective:** To determine the short-term immune response after the SV and AZ vaccinations in HCWs.

**Methods:** In this prospective cohort study, HCWs who completed a 2-dose regimen of the SV or AZ were included. Immune response was evaluated by surrogate viral neutralization test (sVNT) and anti-SARS-CoV-2 total antibody. Blood samples were analyzed at 4 and 12 weeks after the complete SV vaccination and at 4 weeks after each dose of the AZ vaccination. The primary outcome was the seroconversion rate at 4-weeks after complete immunization.

**Results:** Overall, 185 HCWs with a median (IQR) age of 40.5(30.3-55.8) years (94 HCWs in the SV group and 91 in the AZ group) were included. At 4 weeks after completing the SV vaccination, 60.6% (95%CI:50.0-70.6%) had seroconversion evaluated by sVNT(≥68%inhibition), comparable to the patients recovered from mild COVID-19 infection(69.0%), with a rapid reduction to 12.2%(95%CI:6.3-20.8) at 12 weeks. In contrast, 85.7%(95%CI:76.8-92.2%) HCWs who completed the second dose of the AZ for 4 weeks had seroconversion, comparable to the COVID-19 pneumonia patients(92.5%). When using the anti-SAR-CoV-2 total antibody level(≥132 U/ml) criteria, only 71.3% HCWs in the SV group had seroconversion, compared to 100% in the AZ group.

**Conclusion:** A rapid decline of short-term immune response in the HCWs after the SV vaccination indicates the need for a vaccine booster, particularly during the ongoing spreading of the SAR-CoV-2 variants of concern.

## Introduction

With efforts to create immunity against the global spreading of severe acute respiratory syndrome coronavirus 2 (SARS-CoV-2) infection or Coronavirus diseases 2019 (COVID-19), several SARS-CoV-2 vaccines have been developed and widely distributed. In Thailand, inactivated SARS-CoV-2 (CoronaVac®, Sinovac, or SV) and ChAdOx1 nCoV-19 (Vaxzevria®, Oxford-AstraZeneca, or AZ) vaccines have been administered to the front-line health care workers (HCWs) since March 2021. While the SV is an inactivated whole-virion SARS-CoV-2 vaccine, the AZ is a replication-deficient adenoviral vector vaccine.^1, 2^ Therefore, the immunological mechanisms of vaccine-induced protection against SARS-CoV-2 in humans were different.^3^

In general, natural infection creates a protective immunity against re-infection. However, in the COVID-19 patients, the re-infection is possible, whereas the definitive antibody level for protective immunity remains inconclusive.^4, 5^ Moreover, recent evidence suggests that a high neutralizing titer may be required for protection against the circulating SAR-CoV-2 variants of concern (VOCs) infection and symptomatic disease.^6, 7^ To measure the SARS-CoV-2 vaccine-induced protective immune response against the symptomatic infection, the neutralizing antibody titer against the SARS-CoV-2 is a highly predictive indicator. The gold standard for neutralizing antibody detection is a plaque reduction neutralizing test (PRNT_50_), which requires live pathogen and complex laboratory settings. Therefore, the more simplified technique using surrogate virus neutralization test (sVNT), which detects total immunodominant neutralizing antibodies targeting the viral spike (S) protein receptor-binding domain (RBD), is generally used as a high sensitivity and specificity rapid test.^8^ Additionally, the binding antibody (total antibodies including IgG, IgM, and IgA) against the SARS-CoV-2 can be detected with simple laboratory techniques, including rapid test, Enzyme-Linked Immunosorbent Assay (ELISA), and Electrochemiluminescence Assay.

Our study, therefore, aimed to determine the immune response after the SV and AZ vaccination in the HCWs, in comparison with the patients recovered from the COVID-19 using sVNT (%inhibition) and anti-SARS-CoV-2 total antibodies (U/ml). Moreover, the correlation between the sVNT and anti-SARS-CoV-2 total antibodies for estimating vaccine efficacy against SARS-CoV-2 was evaluated.

## Methods

The prospective cohort study included HCWs who received the SARS-CoV2 vaccination at King Chulalongkorn Memorial Hospital, Bangkok, Thailand. The study was approved by the Research Ethics Review Committee, Faculty of Medicine, Chulalongkorn University, and was registered to the Thai Clinical Trial (TCTR20210325003). The protocol was performed in accordance with the declaration of Helsinki and its later amendments. All participants were voluntary to receiving the SARS-CoV-2 vaccination and participating in the study with written informed consent.

### Study Population

Inclusion criteria were as follows: (1) HCWs aged 18 years old and above; and (2) no history of the COVID-19. Patients who had (1) ongoing immunosuppressive medications; (2) any vaccinations within 1 month; or (3) any blood components or intravenous immunoglobulin administrations within 3 months; were excluded. Overall, each vaccination group consisted of 50 participants aged 18-40 years and 50 aged 41-70 years. In the SV group, 0.5 ml (3 μg) of inactivated SARS-CoV-2 (CoronaVac®, Sinovac, or SV) vaccine was administered intramuscularly at the deltoid region, using a 2-dose regimen with an interval of 21-28 days. For the ChAdOx1 nCoV-19 (Vaxzevria®, Oxford-AstraZeneca, or AZ) vaccination group, 0.5 ml of the AZ containing 5×10^10^ viral particles was administered intramuscularly at the deltoid region, using a 2-dose regimen with an interval of 8-10 weeks.

For the COVID-19 group, data were collected from patients diagnosed with the SARS-CoV-2 infection at the Emerging Infectious Diseases Clinical Center at King Chulalongkorn Memorial hospital from March to April 2020, which, at that time, the wild-type SARS-CoV-2 was the main circulating strain in Thailand. The COVID-19 was diagnosed by reverse transcriptase-polymerase chain reaction (RT-PCR) of SARS-CoV-2 from the throat and nasopharyngeal swab samples. The COVID-19 patients were classified according to the clinical severity into mild COVID-19 and COVID-19 pneumonia.

### Demographics and Clinical Data

Baseline demographics and clinical data, including medical history, current medications used, and a history of exposure to the COVID-19 patients, were collected. In addition, for the COVID-19 patients, the clinical characteristics of the SARS-CoV-2 infections were evaluated.

### Sample Collections and Study Protocol

For the vaccination groups, 4-ml clotted blood was collected at baseline for all participants, then at 4 and 12 weeks (+/-2 weeks) after the second dose of the SV vaccination and at 4 weeks (+/-2 weeks) after the first and second doses of the AZ vaccination.

For the COVID-19 group, the blood samples were collected at the time of diagnosis and 4±2 weeks after diagnosis.

### The Detection of Antibody Titers Against SARS-CoV-2

The antibody titers against SARS-CoV-2 were determined by the surrogate neutralizing antibody and total antibodies using the ELISA technique.

#### 1. The SARS-CoV-2 Neutralizing Antibody

The SARS-CoV-2 neutralizing antibody was detected by the blocking ELISA technique of the cPass™ SARS-CoV-2 Neutralization Antibody Detection Kit (GenScript), which the US Food and Drug Administration (FDA) issued Emergency Use Authorizations (EUAs). The protein-protein interaction between Horseradish peroxidase (HRP) conjugated recombinant SARS-CoV-2 RBD fragment (HRP-RBD), and the human ACE2 receptor protein (hACE2) can be blocked by neutralizing antibodies against SARS-CoV-2 RBD. The neutralizing antibody level was detected as percent signal inhibition (%inhibition) following the manufacturer’s protocol. Briefly, serum (negative or positive controls) was diluted 10-fold in a sample diluent (as described in the package insert), mixed with HRP-RBD, then incubated at 37°C for 30 minutes. After incubation, the mixture was added to the hACE2-coated well and incubated at 37°C for 30 minutes. The ELISA plate was then washed with a wash solution for 4 times. The 3,3’,5,5’-tetramethylbenzidine (TMB) substrate was added and incubated for 15 minutes. Finally, the stop solution was added before the optical density (OD) value at 450 nm was read. The antibody level is described as the percentage of signal inhibition or %inhibition.^8^ The percent signal inhibition was calculated using the equation from the manufacturer with the cut-off level for SARS-CoV-2 neutralizing antibody detection of ≥30%inhibition.

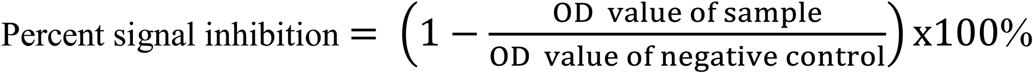

The seroconversion rate was defined as sVNT ≥68%, adopted from the USA-FDA guidance of a high titer of the COVID-19 convalescent plasma.^9^

#### 2. The SARS-CoV-2 Total Antibodies

The SARS-CoV-2 total antibodies were detected by the Elecsys® Anti-SARS-CoV-2 S using Cobas e411 immunoassay analyzers (Roche Diagnostics, Rotkreuz, Switzerland), which is also the US-FDA EUAs. The Elecsys® is the immunoassay for SARS-CoV-2 total antibodies against the RBD of the S antigen detection, and the antibody level is reported as U/ml. Two hundred microliter of serum sample was used following the manufacturer’s protocol. The criteria of negative for anti-SARS-CoV-2-S is <0.8 U/ml. For the participants who had the SARS-CoV-2 total antibodies over the maximum measuring range (which is >250 U/ml), 10-fold diluted samples using the Elecsys® Diluent Universal were re-evaluated. Moreover, the baseline serum of each participant in the vaccination groups was confirmed to be negative for the SARS-CoV-2 total antibodies.^10^ The seroconversion rate was defined as the anti-SARS-CoV-2 total antibodies ≥132 U/ml as suggested by the US-FDA of the high titer for COVID-19 convalescent plasma.^9^

The level of anti-SARS-CoV-2 total antibodies (measured in U/ml) from the Elecsys® test was converted to the BAU/ml following the first WHO International Standard for anti-SARS-CoV-2 immunoglobulin, which 1 U/ml is equivalent to 0.972 BAU/ml.^11^

### Statistical Analysis

Demographics and clinical and laboratory parameters were described in descriptive statistics. Continuous variables were presented as median and interquartile range (IQR). The Wilcoxon rank-sum test was applied to compare the continuous variables between two groups and the Kruskal-Wallis test in case of more than two groups. The Chi-square test or Fisher exact test were used to compare the proportion between groups. The correlation between sVNT and anti-SARS-CoV-2 total antibodies was determined by the Spearman rank test. Statistical significance was considered as P<0.05. STATA version 15.1 (Stata Corp., College Station, Texas) was used for statistical analysis.

## Results

For the vaccination groups, a total of 200 HCWs participated in the study. However, only 185 participants completed the 2-dose regimen of the vaccinations and had the blood collection at 1 month after the second dose (94 and 91 participants in the SV and the AZ groups, respectively). The median (IQR) intervals between the second dose and the blood collection were 23 (22-24) days for the SV group and 19 (16-21) days for the AZ group. For the COVID-19 group, a total of 182 COVID-19 patients were admitted to King Chulalongkorn Memorial Hospital during March and April 2020. One hundred eleven patients had convalescent sera collected at approximately 1 month after diagnosis with a median (IQR) interval of 35 (30-38) days. Of 111 patients, 58 were diagnosed with mild COVID-19 (including upper respiratory tract infection (URI)) and 53 with COVID-19 pneumonia (including 26 moderate and 23 severe pneumonia (who required oxygen supplement or ventilator support) patients). Demographic data and clinical characteristics are described in Table 1. At baseline, the immune response of all participants in the vaccination groups was confirmed to be negative by both tests.

**Table 1:**
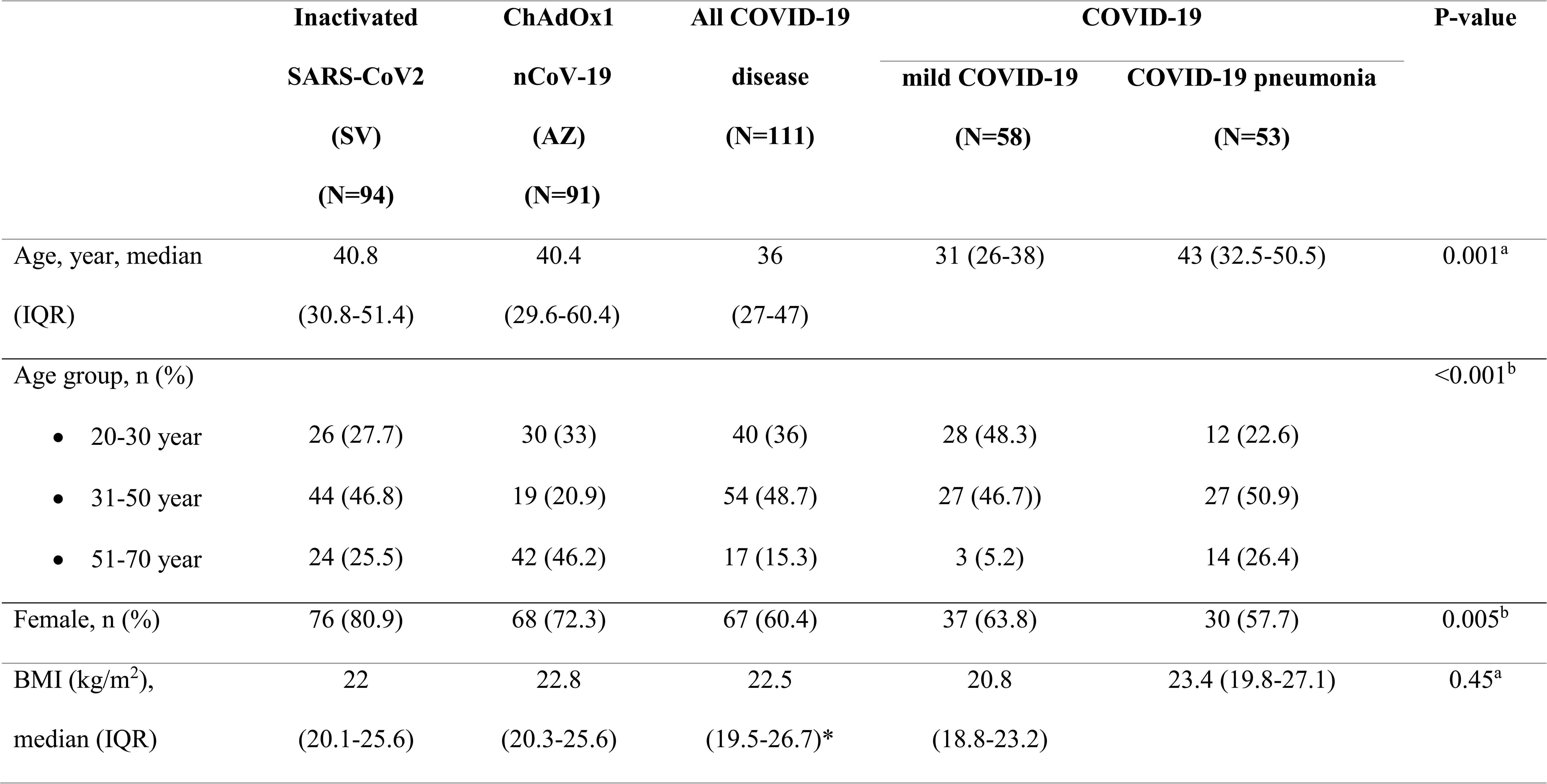
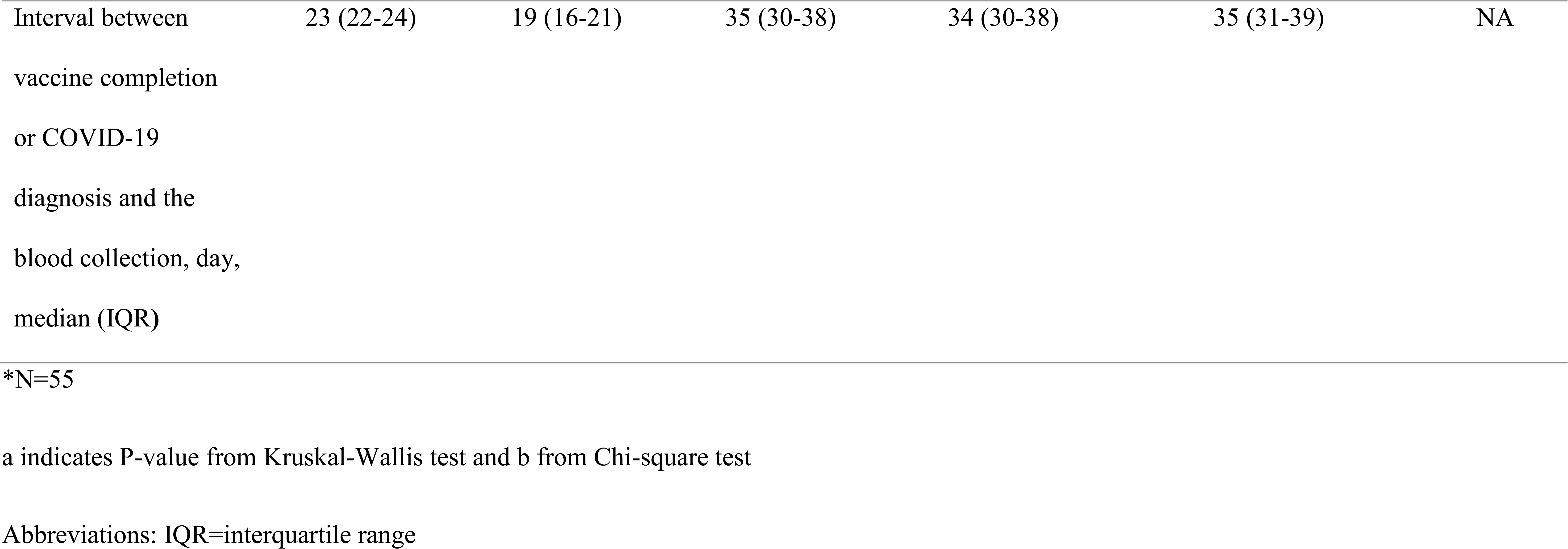
Demographics data of health care workers who received inactivated SARS-CoV-2 vaccine (CoronaVac®, Sinovac, or SV) and ChAdOx1 nCoV-19 vaccine (Vaxzevria®, Oxford-AstraZeneca, or AZ) or patients with COVID-19 diseases

|  | **Inactivated** | **ChAdOx1** | **All COVID-19** | **COVID-19** |  | **P-value** |
| --- | --- | --- | --- | --- | --- | --- |
|  | **SARS-CoV2** | **nCoV-19** | **disease** | **mild COVID-19** | **COVID-19 pneumonia** |  |
|  | **(SV)** | **(AZ)** | **(N=111)** | **(N=58)** | **(N=53)** |  |
|  | **(N=94)** | **(N=91)** |  |  |  |  |
| Age, year, median | 40.8 | 40.4 | 36 | 31 (26-38) | 43 (32.5-50.5) | 0.001<sup>a</sup> |
| (IQR) | (30.8-51.4) | (29.6-60.4) | (27-47) |  |  |  |
| Age group, n (%) |  |  |  |  |  | <0.001<sup>b</sup> |
| • 20-30 year | 26 (27.7) | 30 (33) | 40 (36) | 28 (48.3) | 12 (22.6) |  |
| • 31-50 year | 44 (46.8) | 19 (20.9) | 54 (48.7) | 27 (46.7)) | 27 (50.9) |  |
| • 51-70 year | 24 (25.5) | 42 (46.2) | 17 (15.3) | 3 (5.2) | 14 (26.4) |  |
| Female, n (%) | 76 (80.9) | 68 (72.3) | 67 (60.4) | 37 (63.8) | 30 (57.7) | 0.005<sup>b</sup> |
| BMI (kg/m<sup>2</sup>), | 22 | 22.8 | 22.5 | 20.8 | 23.4 (19.8-27.1) | 0.45<sup>a</sup> |
| median (IQR) | (20.1-25.6) | (20.3-25.6) | (19.5-26.7)* | (18.8-23.2) |  |  |
| Interval between<br>vaccine completion<br>or COVID-19<br>diagnosis and the<br>blood collection, day,<br>median (IQR) | 23 (22-24) | 19 (16-21) | 35 (30-38) | 34 (30-38) | 35 (31-39) | NA |

\*N=55
a indicates P-value from Kruskal-Wallis test and b from Chi-square test
Abbreviations: IQR=interquartile range

### The SARS-CoV-2 Neutralizing Antibodies (sVNT, %inhibition)

The sVNTs (%inhibition) at 4 weeks after completing the second dose of the vaccinations are demonstrated in Table 2 and Figure 1. The median (IQR) of SARS-CoV-2 sVNT was 77.0 (58.5-87.9) %inhibition in the SV group, 90.4 (75.2-96.2) %inhibition in the AZ group, 80.8 (61.5-92.1) %inhibition in the mild COVID-19 group, and 94.5 (88.1-95.8) %inhibition in the COVID-19 pneumonia group.

**Figure 1:**
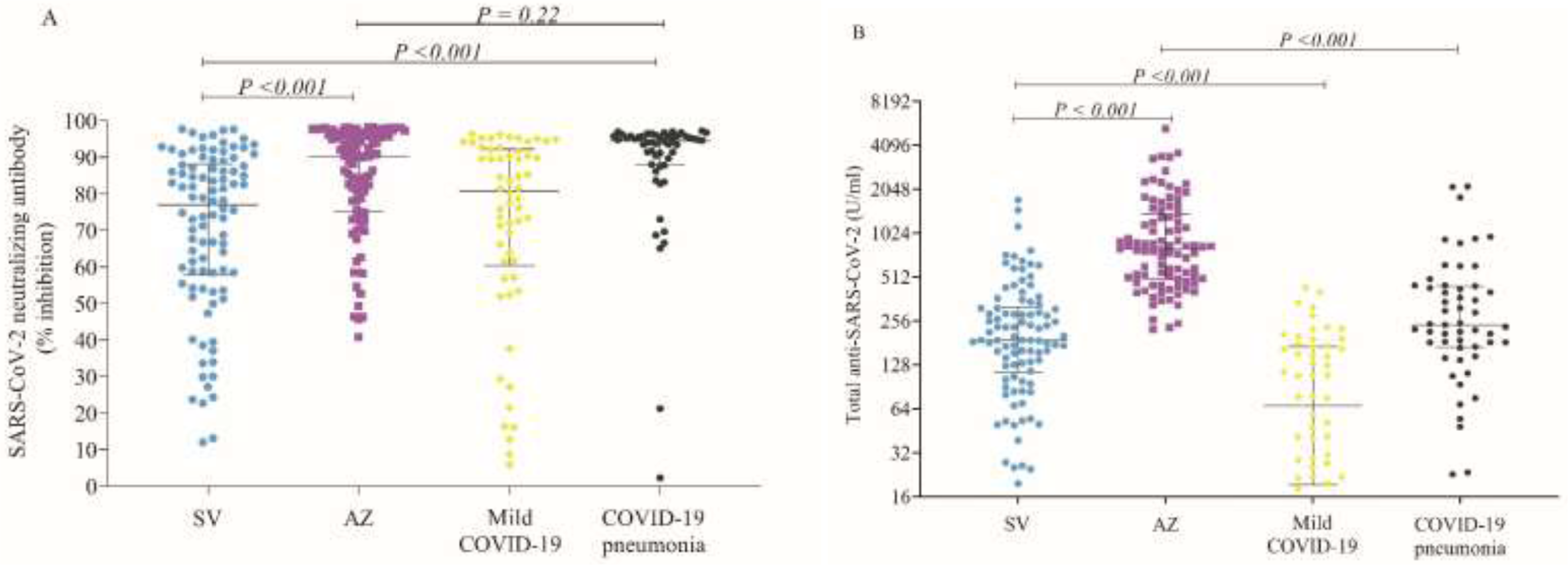
A. the SARS-CoV-2 Neutralizing antibody using the cPass^TM^ test (% inhibition) B. the anti-SARS-CoV-2 total antibody using the Elecys^TM^ test (U/mL) after complete 2-dose of inactivated SARS-CoV-2 vaccine (CoronaVac®, Sinovac, or SV) and ChAdOx1 nCoV-19 vaccine (Vaxzevria®, Oxford-AstraZeneca, or AZ) compare to convalescent sera after COVID-19 diseases

**Table 2:**
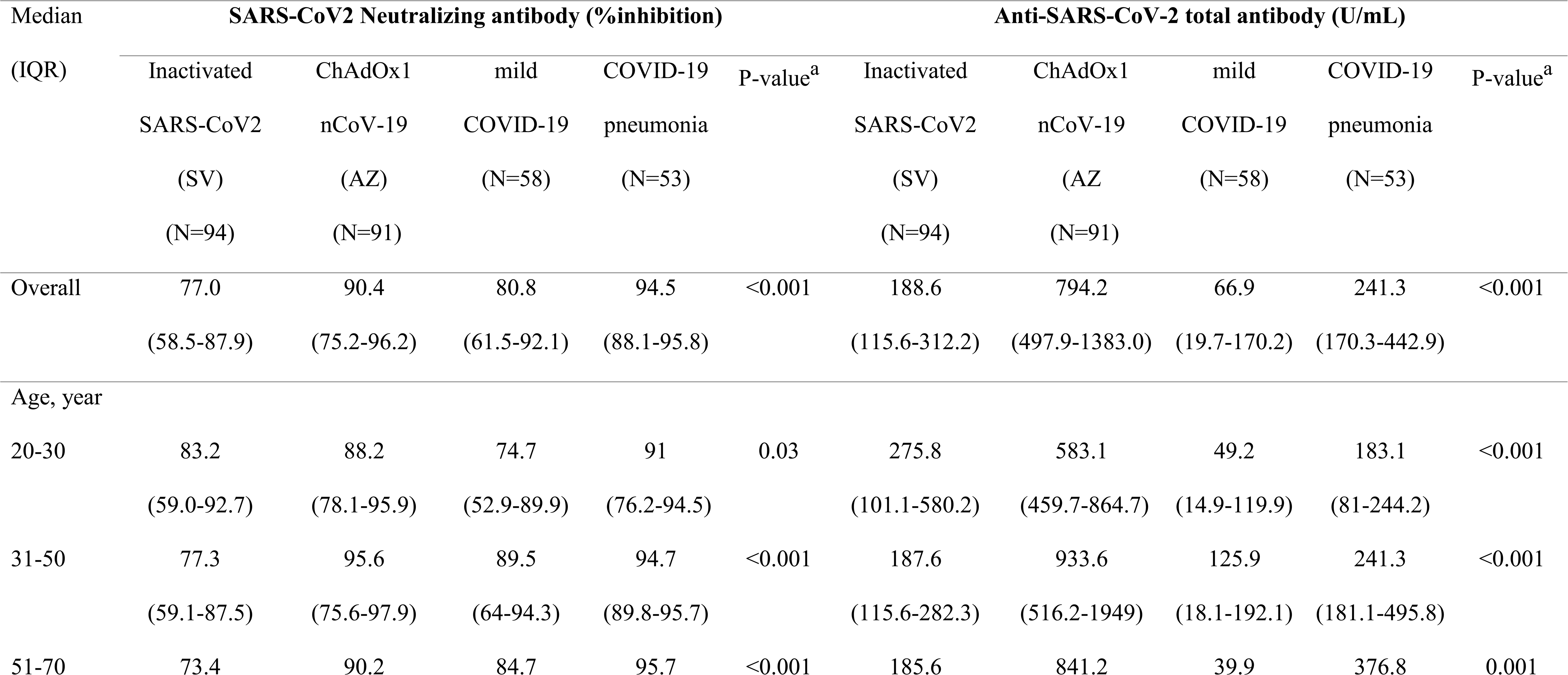
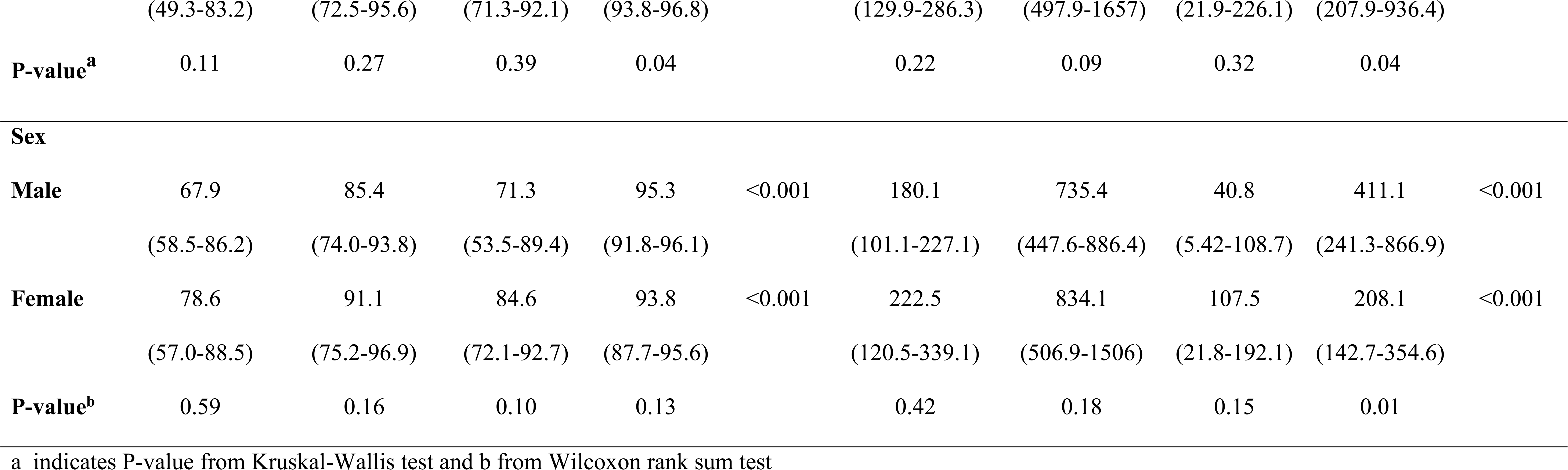
Median (IQR) of the SARS-CoV2 Neutralizing antibody using the cPass^TM^ test and the anti-SARS-CoV-2 total antibody using the Elecys^TM^ test at 4 weeks after complete 2-dose of inactivated SARS-CoV-2 vaccine (CoronaVac®, Sinovac, or SV) and ChAdOx1 nCoV-19 vaccine (Vaxzevria®, Oxford-AstraZeneca, or AZ) compare to convalescent sera after COVID-19 disease

For the SV group, sVNT tended to be lower in the older age group but was not statistically significant (83.2%inhibition in the 20-30 years group vs 73.4%inhibition in the 51-60 years group, P=0.18). There was no statistically significant difference of sVNT between age ranges and sexes in each group. (Table 2) Comparing to the convalescent sera of the COVID-19 patients, the SV group had a level of SARS-CoV-2 neutralizing antibodies comparable to the mild COVID-19 group. In contrast, the AZ group had a higher level of neutralizing antibodies comparable to the COVID-19 pneumonia group. (Figure 1)

At 12 weeks after the second dose of the SV vaccination, the median (IQR) of SARS-CoV-2 sVNT was 38.7 (22.1-55.7) %inhibition, which significantly decreased from the immune response at 4 weeks (77.0 (58.5-87.9) %inhibition) (P<0.001). (Figure 2A, Supplementary Table 1) On the contrary, the median (IQR) of SARS-CoV-2 sVNT at 4 weeks after the first dose of AZ was 37.6 (6.5-63.2) %inhibition and significantly increased after the second dose (P<0.001). (Figure 2B, Supplementary Table 2)

**Figure 2:**
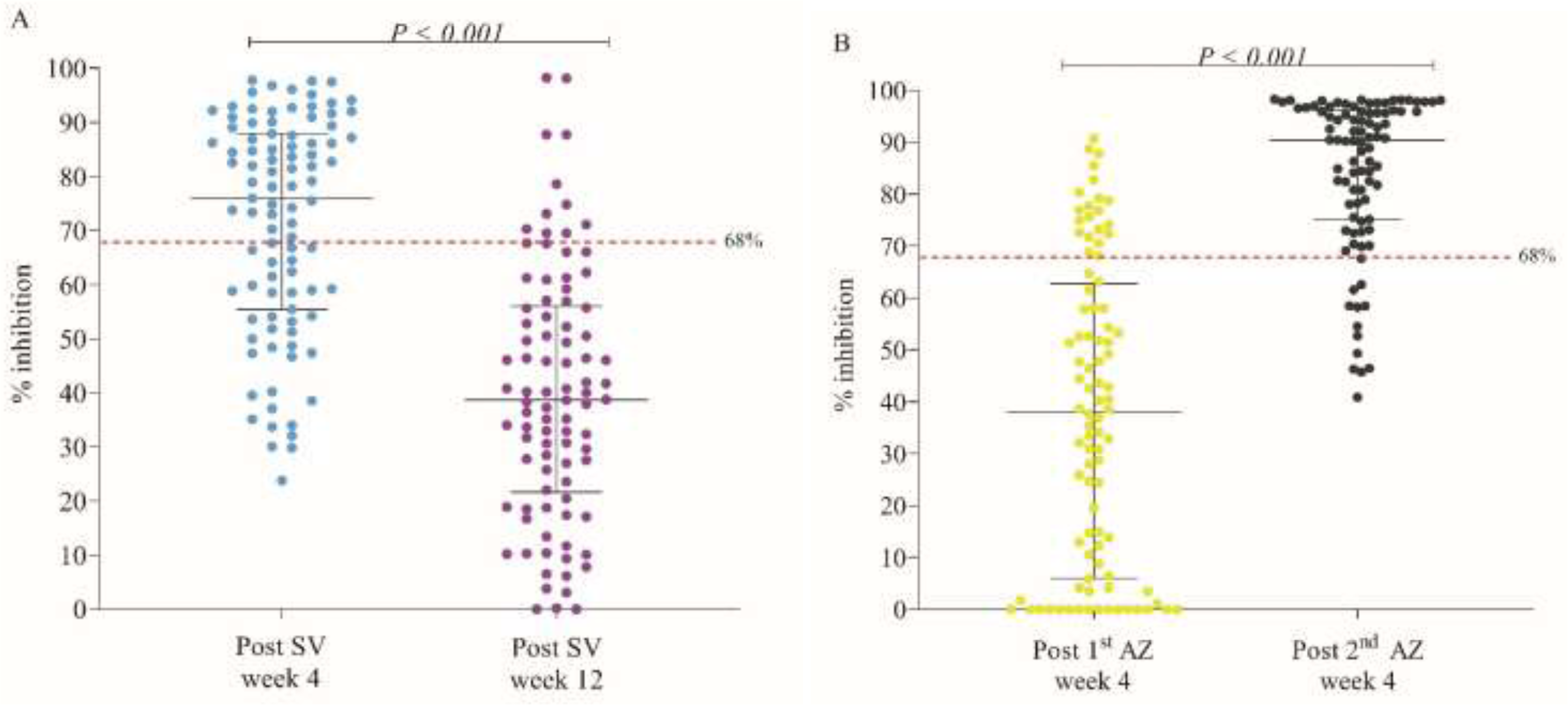
A. The kinetic of the SARS-CoV-2 Neutralizing antibody using the cPass^TM^ test (% inhibition) in health care workers who received inactivated SARS-CoV-2 vaccine (CoronaVac®, Sinovac, or SV) at 4 and 12 weeks after complete 2-dose immunization. B. in health care workers who received ChAdOx1 nCoV-19 vaccine (Vaxzevria®, Oxford-AstraZeneca, or AZ) at 4 weeks after the first and the second dose of immunization

For the US-FDA guidance of a high titer of the COVID-19 convalescent plasma, the %inhibition using the cPass^TM^ SARS-CoV-2 Neutralization Antibody Detection Kit should be 68% and over (or ≥68%inhibition). At 4 weeks after completing the vaccinations, 57/94 (60.6%) participants in the SV group had ≥68%inhibition, compared to 78/91 (85.7%) participants in the AZ group. (Table 3) However, only 11/90 (12.2%) participants in the SV group had the high titers at 12 weeks after the second dose. (Supplementary Table 1). For the AZ group, 18/87 (20.7%) participants had ≥68%inhibition at 4 weeks after the first dose with the median (IQR) sVNT of 37.6 (6.5-63.2) %inhibition. (Supplementary Table 2)

**Table 3:** The seroconversion rates of health care workers who received inactivated SARS-CoV-2 vaccine (CoronaVac®, Sinovac, or SV) and ChAdOx1 nCoV-19 vaccine (Vaxzevria®, Oxford-AstraZeneca, or AZ) and patients with COVID-19 infection who achieved the SARS-CoV2 neutralizing antibody ≥68%inhibition and ≥80%inhibition and anti-SARS-CoV-2 total antibody of ≥132 U/mL

| Seroconversion rate | Inactivated SARS-CoV2<br>(SV)<br>(N=94) | ChAdOx1 nCoV-19<br>(AZ)<br>(N=91) | Mild COVID-19<br>(N=58) | COVID-19<br>Pneumonia<br>(N=53) | P-value |
| --- | --- | --- | --- | --- | --- |
| %Inhibition $\geq 68\%$ (N, (%)) | 57 (60.6) | 78 (85.7) | 40 (69) | 49 (92.5) | <0.001 |
| %Inhibition $\geq 80\%$ (N, (%)) | 43 (45.7) | 64 (70.3) | 30 (51.7) | 46 (86.8) | <0.001 |
| SARS-CoV-2 total antibody<br>$\geq 132$ U/mL (N, (%)) | 67 (71.3) | 91 (100) | 20 (34.5) | 44 (83) | <0.001 |
| P-value from Chi-square test |  |  |  |  |  |

For sensitivity analysis, a higher cPass^TM^ SARS-CoV-2 Neutralization Antibody Detection Assays of ≥80%inhibition was used as a surrogate marker for high sVNT against the SARS-CoV-2 variants of concerns. Consequently, only 43/94 (45.7%) participants in the SV group and 64/91 (70.3%) participants in the AZ group had ≥80%inhibition, compared to 46/53 (86.8%) patients in the COVID-19 pneumonia group (P<0.001). (Table 3)

### The SARS-CoV-2 Total Antibodies

The median (IQR) of anti-SARS-CoV-2 total antibodies were 188.6 (115.6-312.2) U/ml in the SV group, 794.2 (497.9-1383.0) U/ml in the AZ group, 66.9 (19.7-170.2) U/ml in the mild COVID-19 group, and 794.2 (497.9-1383) U/ ml in the COVID-19 pneumonia group. In the SV group, the anti-SARS-CoV-2 total antibodies among the older age group tended to be decreased but were not statistically significant (275.8 U/ml in the 20-30 years group vs 185.6 U/ml in the 51-60 years group, P=0.28).

Using the US-FDA guidance of a high titer of the COVID-19 convalescent plasma, the cut-off index (COI) for the Elecys^TM^ test anti-SARS-CoV-2 total antibodies is ≥132 U/ ml. Using the US-FDA COI criteria, 67/94 (71.3%) participants in the SV group and 91/91(100%) participants in the AZ group had seroconversion. (Table 3) Notably, the anti-SARS-CoV-2 total antibodies of the AZ group were significantly higher than the COVID-19 pneumonia group (P<0.001). (Figure 1)

According to the generalized additive models to estimate the immune marker values, in this case, the anti-SARS-CoV-2 total antibodies using the Elecys^TM^ kit technique (converted to BAU/ml), the vaccine efficacy of SV and AZ were 50% and 70%, respectively.^11^ (Supplementary Table 3, 4)

### Correlation Between The SARS-CoV-2 Neutralizing Antibodies and The Anti-SARS-CoV-2 Total Antibodies

The Spearman correlation coefficient between the sVNT and the anti-SARS-CoV-2 total antibodies was 0.79 for the SV group and 0.86 for the AZ group. (Figure 3) Interestingly, 78/91 (86.7%) participants in the AZ group had both high neutralizing antibodies (sVNT ≥68%inhibition) and high binding antibodies (anti-SARS-CoV-2 total antibodies ≥132 U/ml), compared to 55/94 (58.5%) participants in the SV group (P<0.001).

**Figure 3:**
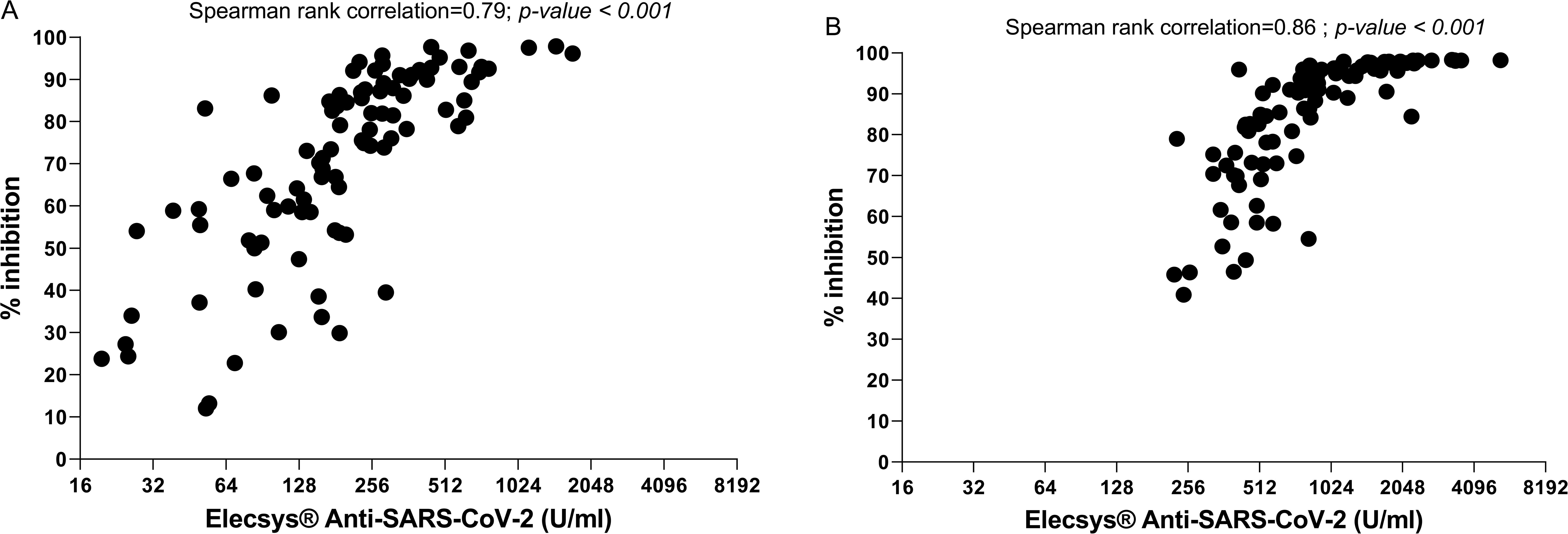
Correlation between the SARS-CoV-2 neutralizing antibody using the cPass^TM^ test (% inhibition) and the anti-SARS-CoV-2 total antibody using the Elecys^TM^ test (U/mL) in health care workers who received inactivated SARS-CoV-2 vaccine (CoronaVac®, Sinovac, or SV) (A) and ChAdOx1 nCoV-19 vaccine (Vaxzevria®, Oxford-AstraZeneca, or AZ) (B) vaccination.

## Discussion

This study presented the short-term immune response of Thai HCWs after completing the 2-dose regimens of both the SV and AZ for one month and at three months after the SV vaccination. In this study, the surrogate immune markers were used to predict the vaccine efficacy to protect against the SARS-CoV-2 infection and/or symptomatic disease.

The immune response of the AZ group as measured by the sVNT was higher than the SV group (85.7% vs 60.8%inhibition). Additionally, the binding antibodies, or the anti-SARS-CoV-2 total antibodies, of ≥132 IU/ml was 100% in the AZ group and only 71% in the SV group. At 3-month after completing the SV vaccination, sVNT significantly declined to 12.2%. Similarly, the rapid decline of protective immunity was observed after the SARS-CoV-2 infection, as described by the progressive decline of the sVNT after 5-8 weeks post-infection and the decreased systemic IgA antibody level at 8 weeks post-infection.^12–13^ In our study, HCWs who completed the 2-dose regimen of the SV vaccination, demonstrated a rapid decrease of sVNT at 12 weeks afterwards, and the anti-SARS-CoV-2 total antibodies were just equivalent to the mild COVID-19 patients. As the SARS-CoV-2 neutralizing antibodies level implicated the vaccine efficacy against the SARS-CoV-2 infection and symptomatic disease, the rapid reduction of the sVNT should prompt the concerns of reduced vaccine efficacy at 1-6 months after the complete SV vaccination. Moreover, the protective immunity against the SARS-CoV-2 VOC tends to require a high sVNT, though the definitive level is inconclusive.

Thus, HCWS who previously received the SV vaccination should have a vaccine booster to accelerate the sVNT level. In the AZ group, though the immune response was low after the first dose, then dramatically increase after the second dose. Therefore, the interval period of the vaccination should be shortened to accelerate the sVNT level to protect against the SARS-CoV-2 VOC earlier.

The study in Chile reported the vaccine effectiveness of the SV to be 65.9% for the prevention of COVID-19.^14^ In our study, the seroconversion rate was an immune marker that implicated the protective immunity against the SARS-CoV-2 infection. Therefore, the 60.8% seroconversion rate in the SV group should be equivalent to the reported vaccine effectiveness from Chile. Notably, the study in Chile was conducted during Feb-April 2021, when the circulating strains were Alpha (B.117) and Gamma (P1). Furthermore, in the SV group, the immune response tended to decrease with age, corresponding with the data from the phase 1/2 clinical trials of the SV in healthy adults aged 60 years and older.^15^ This finding supports the Thai government initial policy of vaccine allocation, provide the SV to younger age group (18-59 years) and the AZ to the elderly (≥60 years).

In the AZ group, a robust immune response was observed in all age groups, including the elderly, corresponding with the phase 2/3 study of the ChAdOx1 nCoV-19 vaccine in young and old adults.^16^ In addition, vaccine efficacy of the AZ was similar to the study in Brazil, South Africa, and the UK.^17^ Of note, the level of the AZ-induced immune response was 4-time higher than the convalescent sera after the wild-type SARS-CoV-2 infection. However, vaccine efficacy was expected to be reduced for the SARS-CoV-2 VOC. In Thailand, B1.617.2 (Delta) variant has been the main circulating SARS-CoV-2 VOC since July 2021. According to the study from the serum of the mRNA-1273 (Spikevax®, Moderna) vaccinated patients, the neutralizing antibody titers against Delta was 2.9 times less susceptible to neutralization^18^; therefore, a booster vaccine may also require after the complete AZ vaccination. The Com-Cov study also supported the new vaccine regimen of heterogenous prime-boost COVID-19 vaccination using AZ prime and boost with mRNA-BNT162b2 (Comirnaty®, Pfizer–BioNTech).^19^

A single dose of AZ also demonstrated efficacy in reducing the disease severity. Regarding the immunological mechanism of the AZ vaccine-induced protection against SARS-CoV-2, the low sVNT level after the first dose of AZ may be explained by the sole measurement of humoral response mainly from the B-cell activation. However, as a viral vector vaccine, the cellular responses (including the T cells) were also activated but were under-estimated by the sVNTs. Thus, the AZ-induced Th1 and CD8 T cell activity after the first dose of the AZ may contribute to the vaccine efficacy in ameliorating the disease severity.^3, 20^

Moreover, a correlation coefficient between sVNT and anti-SARS-CoV-2 total antibodies were higher in the AZ group (0.86) than the SV group (0.79). The lower correlation coefficient may be explained by the different immunological mechanisms of the inactivated viral vaccine, including the SV, which induces a lower neutralising antibody level. Consequently, though some individuals had high titers of the SARS-CoV-2 total antibodies, the sVNT was low, indicating that the majority of anti-SARS-CoV-2 total antibodies were not neutralizing antibodies. Although other non-neutralizing antibodies might also contribute to the protective immunity,^3^ the neutralization level is highly predictive of immune protection.^21^ Therefore, the anti-SARS-CoV-2 total antibodies following the SV vaccination might not sufficiently represent the immune response against the SARS-CoV-2 infection, using the more accessible laboratory technique.

There were several strengths of this study. Firstly, as the HCWs were the first group who received the SARS-CoV-2 vaccinations in Thailand, our data will guide the policymakers of the Thai National Vaccine Roll-Out Program to outline the vaccine programs for the country. Secondly, the HCWs has an effective surveillance protocol to screen for the SAR-CoV-2 infection. Therefore, if no episode of SARS-CoV-2 infection was documented, the immune response should solely result from the vaccination.

This study has some limitations. Firstly, there were no definitive cut-off level of the sVNT and the SARS-CoV-2 total antibodies for protection against both wild-type SARS-CoV-2 virus and the SARS-CoV-2 VOC. Therefore, a high sVNT ≥68% and 80%inhibition by the US-FDA were adopted to implicate the protective immunity. Moreover, previous studies reported the high correlation between the sVNT and the PRNT_50_ and the equivalent sVNTs of the convalescent sera of the severe COVID-19 patients. Secondly, both sVNT and the SARS-CoV-2 total antibodies only represented the humoral immune response. Further studies on the cellular immune responses after the vaccination are ongoing, comparing the T-cell activities by the ELISpot assay between the individuals who received the AZ and the SV vaccinations. Theoretically, the AZ vaccine, a viral vector vaccine platform, should have a better cell-mediated immune response than the SV vaccine, an inactivated virus vaccine platform.

In general, a highly effective vaccine with the lowest possible side effects should be offered to the front-line HCWs. From our data, the HCWs who completed 2 doses of the SV vaccination should be considered for a booster vaccine, and the HCWs who received the first dose of the AZ vaccination should shorten the interval for the second dose.

## Conclusion

During the COVID-19 pandemic, the HCWs are at very high risk, as they are the front-line team to deal with the patients. Therefore, all protective measures, including non-pharmacological (including personal protective equipment) and pharmacological (including a SARS-CoV-2 vaccination), should be highly regarded. Currently, high proportions of young HCWs in Thailand completed the SV vaccinations. However, after completing the vaccination, the short-term immune response appeared to be lower in the SV group than in the AZ group. Moreover, the HCWs in the SV group experienced a rapid decline of the immune response as early as three months after the complete vaccination. In this study, our results emphasized the urgent need for the booster vaccine for the HCWs, particularly in the SV vaccination, in amidst the circulating SARS-CoV-2 VOC such as the delta strain.

## Data Availability

The data that support the findings of this study are available on request from the corresponding author, [OP]. The data are not publicly available due to their containing information that could compromise the privacy of research participants.

## Acknowledgements

We would like to thank all health care workers at King Chulalongkorn Memorial Hospital (KCMH) as well as the study team including Saithip khumpiwdum, Phattharasuda Yodbutdee and Thitima maneepornpol. We also thank Susama Chokesuwattanaskul, of the Department of Ophthalmology, Faculty of Medicine, Chulalongkorn University and King Chulalongkorn Memorial Hospital, for help with manuscript editing.

## Author contributions

TP, SW, NH, LP GP, and OP contributed substantially to the conception and design of this study. WJ, NC, RP and PT contributed substantially to the acquisition of the data. WJ, NP, SW, VR, NH, TP and JS analyzed and interpreted the data. WJ, NP, and TP drafted the manuscript. LP, GS, TP, SW, NH and OP contributed substantially to the manuscript revision. All the authors approved the final version submitted for publication and take responsibility for the statements made in the published article.

---

**Supplement Table 1.** The median (IQR) of the SARS-CoV-2 neutralizing antibody using the cPass^TM^ test (% inhibition) in health care workers who received inactivated SARS-CoV-2 vaccine (CoronaVac®, Sinovac, or SV) at 4-week and 12-week after complete immunization.

| **SARS-CoV-2 Neutralizing antibody using the cPass™ test (%inhibition)** |  |  |  |
| --- | --- | --- | --- |
| **Median, (IQR)** | **Weeks 4** | **Weeks 12** | **P-value<sup>a</sup>** |
|  | **N=94** | **N=90** |  |
| **Overall** | 77.0 (58.5-87.9) | 38.7 (22.1-55.7) | <0.001 |
| **Age, year** |  |  |  |
| **20-30** | 83.2 (59.0-92.7) | 40 (31.7-66) | <0.001 |
| **31-50** | 77.3 (59.1-87.5) | 43.7 (21.2-57.4) | <0.001 |
| **51-70** | 73.4 (49.3-83.2) | 32.4(15.3-43.3) | <0.001 |
| **P-value<sup>b</sup>** | 0.11 | 0.06 |  |
| **Sex** |  |  |  |
| **Male** | 67.9 (58.5-86.2) | 37 (15.1-58.3) | 0.006 |
| **Female** | 78.6 (57.0-88.5) | 38.8 (27.6-55.8) | <0.001 |
| **P-value<sup>c</sup>** | 0.59 | 0.85 |  |

a indicates P-value from Wilcoxon sign rank test, b from Kruskal-Wallis test and c from Wilcoxon rank sum test

**Supplement Table 2.**
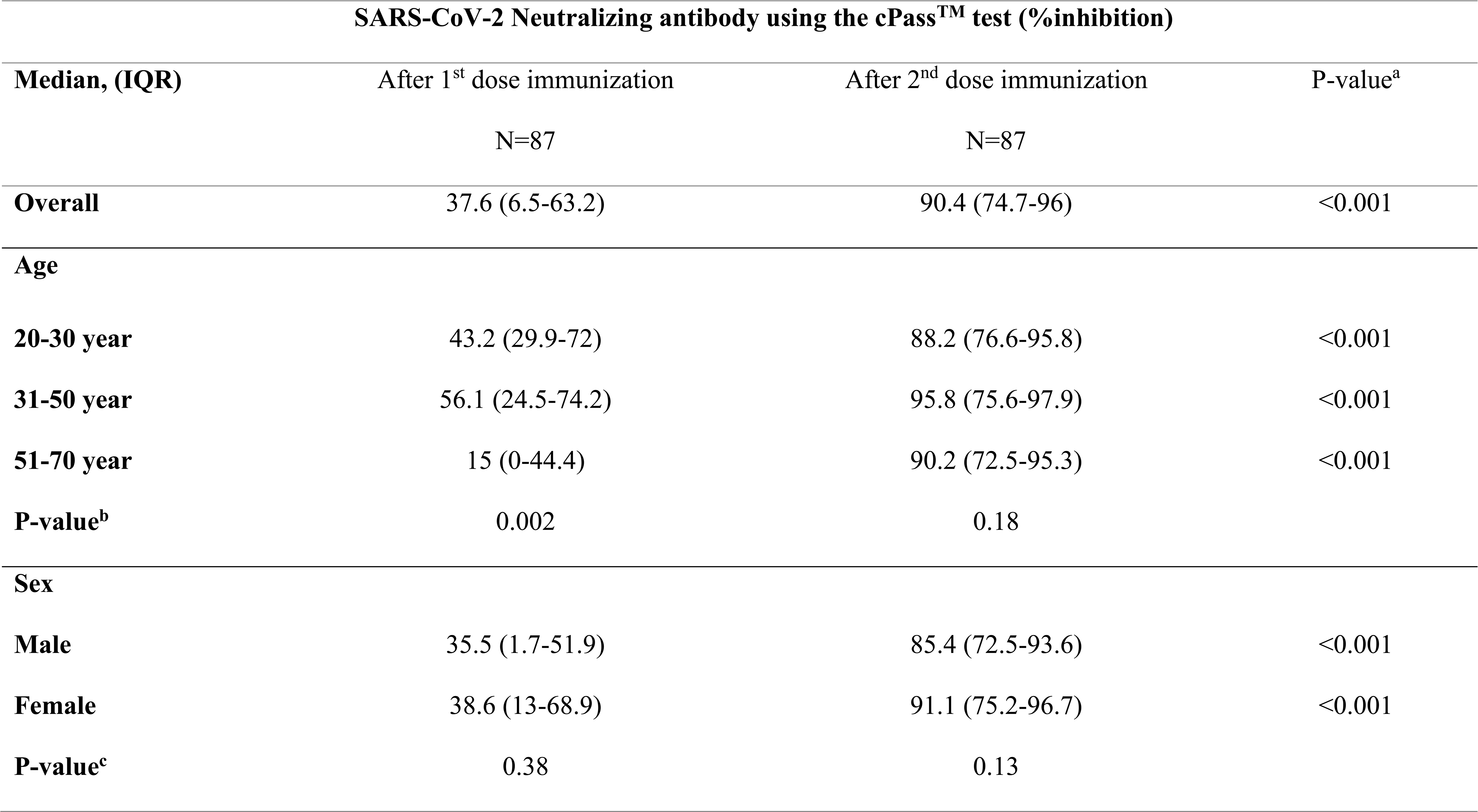
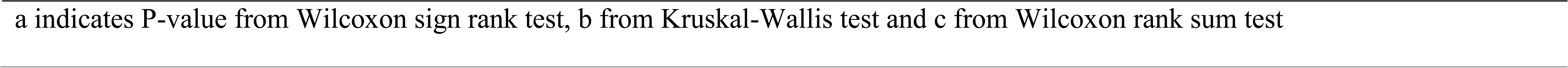
The median (IQR) of the SARS-CoV-2 neutralizing antibody using the cPass^TM^ test (% inhibition) in health care workers who received ChAdOx1 nCoV-19 vaccine (Vaxzevria®, Oxford-AstraZeneca, or AZ) at 4-week and 12-week after the 1^st^ and 2^nd^ dose immunization.

**Supplementary Table 3:**
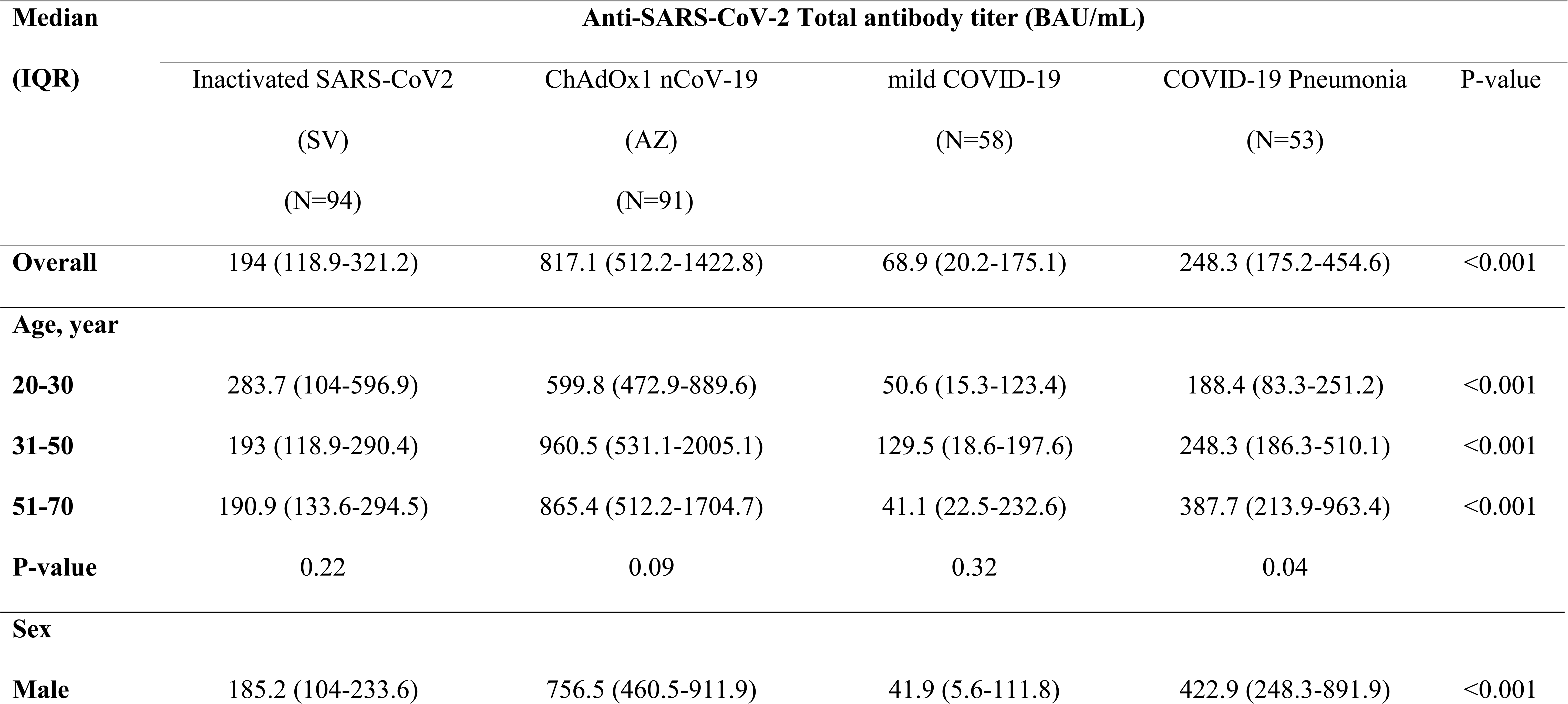
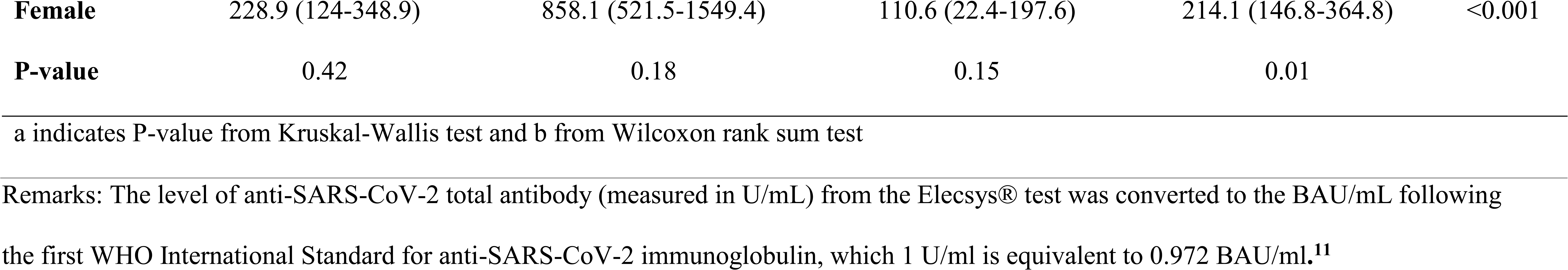
Median (IQR) of the anti-SARS-CoV-2 total antibody (BAU/mL) using the Elecys^TM^ test at 4 weeks after complete 2-dose of inactivated SARS-CoV-2 vaccine (CoronaVac®, Sinovac, or SV) and ChAdOx1 nCoV-19 vaccine (Vaxzevria®, Oxford-AstraZeneca, or AZ) compare to compare to convalescent sera after COVID-19 disease

**Supplementary Table 4:** Vaccine efficacy according to the generalized additive models with immune marker values, which is anti-SARS-CoV-2 total antibody (BAU/mL) using the Elecys^TM^ kit technique^9^ in health care workers after complete 2-dose of inactivated SARS-CoV-2 vaccine (CoronaVac®, Sinovac, or SV) and ChAdOx1 nCoV-19 vaccine (Vaxzevria®, Oxford-AstraZeneca, or AZ) compare to compare to convalescent sera after COVID-19 disease

| Vaccine Efficacy | Anti-SARS-CoV-2 total antibody (BAU/mL) |  |  |  |  |
| --- | --- | --- | --- | --- | --- |
|  | Inactivated SARS-CoV2<br>(SV)<br>(N=94) | ChAdOx1 nCoV-19<br>(AZ)<br>(N=91) | mild COVID-19<br>(N=58) | COVID-19<br>pneumonia<br>(N=53) | P-value |
| **50% (≥18 BAU/mL)** | 94 (100) | 91 (100) | 45 (77.6) | 53 (100) | <0.001 |
| **60% (≥50 BAU/mL)** | 88 (93.6) | 91 (100) | 31 (53.5) | 50 (94.3) | <0.001 |
| **70% (≥165 BAU/mL)** | 58 (61.7) | 91 (100) | 17 (29.3) | 41 (77.4) | <0.001 |
| **80% (≥506 BAU/mL)** | 13 (13.8) | 69 (75.8) | 0 | 11 (20.8) | <0.001 |
| **90% (≥2360 BAU/mL)** | 0 | 7 (7.7) | 0 | 0 | 0.001 |

## Reference

1. Folegatti PM, Ewer KJ, Aley PK, Angus B, Becker S, Belij-Rammerstorfer S, et al. Safety and immunogenicity of the ChAdOx1 nCoV-19 vaccine against SARS-CoV-2: a preliminary report of a phase 1/2, single-blind, randomised controlled trial. Lancet. 2020;396:467–78.

2. Zhang Y, Zeng G, Pan H, Li C, Hu Y, Chu K, et al. Safety, tolerability, and immunogenicity of an inactivated SARS-CoV-2 vaccine in healthy adults aged 18-59 years: a randomised, double-blind, placebo-controlled, phase 1/2 clinical trial. Lancet Infect Dis. 2021;21:181–92.

3. Sadarangani M, Marchant A, Kollmann TR. Immunological mechanisms of vaccine-induced protection against COVID-19 in humans. Nat Rev Immunol. 2021:1–10.

4. Wang J, Kaperak C, Sato T, Sakuraba. COVID-19 reinfection: a rapid systematic review of case reports and case series. J Investig Med. 2021;69:1253–5.

5. Lumley SF, O’Donnell D, Stoesser NE, Matthews PC, Howarth A, Hatch SB, et al. Antibody Status and Incidence of SARS-CoV-2 Infection in Health Care Workers. N Engl J Med. 2021;384:533–40.

6. Abu-Raddad LJ, Chemaitelly H, Butt AA. Effectiveness of the BNT162b2 Covid-19 vaccine against the B.1.1.7 and B.1.351 variants. N Engl J Med. 2021;385:187–9.

7. Bernal JL, Andrew N, Gower C, Gallaher E, Simmons R, et al. Effectiveness of COvid-19 vaccines against the B.1.617.2 (delta) variant. N Engl J Med. 2021; Jul 21 doi: 10.1056/NEJMoa2108891. [Epub ahead of print].

8. Tan CW, Chia WN, Qin X, Liu P, Chen MI, Tiu C, et al. A SARS-CoV-2 surrogate virus neutralization test based on antibody-mediated blockage of ACE2-spike protein-protein interaction. Nat Biotechnol. 2020;38:1073–8.

9. De Santis GC, Mendrone A, Langhi D Jr, Covas DT, Fabron A Jr, Cortez AJP, et al. Suggested guidelines for convalescent plasma therapy for the treatment of COVID-19. Hematol Transfus Cell Ther. 2021;43:212–3.

10. Elecsys Anti-SARS-CoV-2. [cited 2021 Jul 16]. Available from: https://www.fda.gov/media/137605/download

11. . Feng S, Phillips DJ, White T, Sayal H, Aley PK, Bibi S, et al. Correlates of protection against symptomatic and asymptomatic SARS-CoV-2 infection. medRxiv 21258528 [Preprint]. 2021 [cited 2021 Jul 10]. Available from: https://doi.org/10.1101/2021.06.21.21258528

12. Marot S, Malet I, Leducq V, Zafilaza K, Sterlin D, Planas D, et al. Rapid decline of neutralizing antibodies against SARS-CoV-2 among infected healthcare workers. Nat Commun. 2021;12:844.

13. Dispinseri S, Secchi M, Pirillo MF, Tolazzi M, Borghi M, Brigatti C, et al. Neutralizing antibody responses to SARS-CoV-2 in symptomatic COVID-19 is persistent and critical for survival. Nat Commun. 2021;12:2670.

14. Jara A, Undurraga EA, González C, Paredes F, Fontecilla T, Jara G, et al. Effectiveness of an Inactivated SARS-CoV-2 Vaccine in Chile. N Engl J Med. 2021; Jul 7. doi: 10.1056/NEJMoa2107715. [Epub ahead of print].

15. Iversen PL, Bavari S. Inactivated COVID-19 vaccines to make a global impact. Lancet Infect Dis. 2021;21:746–8.

16. Ramasamy MN, Minassian AM, Ewer KJ, Flaxman AL, Folegatti PM, Owens DR, et al. Safety and immunogenicity of ChAdOx1 nCoV-19 vaccine administered in a prime-boost regimen in young and old adults (COV002): a single-blind, randomised, controlled, phase 2/3 trial. Lancet. 2021;396:1979–93.

17. Voysey M, Clemens SAC, Madhi SA, Weckx LY, Folegatti PM, Aley PK, et al. Safety and efficacy of the ChAdOx1 nCoV-19 vaccine (AZD1222) against SARS-CoV-2: an interim analysis of four randomised controlled trials in Brazil, South Africa, and the UK. Lancet. 2021;397:99–111.

18. Edara VV, Pinsky BA, Suthar MS, Lai L, Davis-Gardner ME, Floyd K, et al. Infection and Vaccine-Induced Neutralizing-Antibody Responses to the SARS-CoV-2 B.1.617 Variants. N Engl J Med. 2021; Jul 7. doi: 10.1056/NEJMc2107799. [Epub ahead of print].

19. Shaw RH, Stuart A, Greenland M, Liu X, Nguyen Van-Tam JS, Snape MD, et al. Heterologous prime-boost COVID-19 vaccination: initial reactogenicity data. Lancet. 2021;397:2043–6.

20. Ewer KJ, Barrett JR, Belij-Rammerstorfer S, Sharpe H, Makinson R, Morter R, et al. T cell and antibody responses induced by a single dose of ChAdOx1 nCoV-19 (AZD1222) vaccine in a phase 1/2 clinical trial. Nat Med. 2021;27:270–8.

21. Khoury DS, Cromer D, Reynaldi A, Schlub TE, Wheatley AK, Juno JA, et al. Neutralizing antibody levels are highly predictive of immune protection from symptomatic SARS-CoV-2 infection. Nat Med. 2021;27:1205–11.

